# Intravenous rehydration in children with severe malnutrition: a systematic review and meta-analysis

**DOI:** 10.1101/2025.08.18.25333854

**Authors:** J.E. Dewez, T. Sunyoto, C. Mogaka, M.E. Coldiron, H.A. Sainna, S. Ouattara, R. Petrucci, E.C. George, K. Maitland

## Abstract

**Background:** The comparative efficacy and safety of intravenous rehydration (IVR) and oral rehydration (OR) strategies in children with severe acute malnutrition (SAM) remains uncertain.

**Methods:** We systematically reviewed randomized clinical trials (RCTs) comparing the use of IVR and oral rehydration (standard of care) in children with SAM hospitalized with severe dehydration secondary to gastroenteritis. The primary outcome was in-hospital mortality. Secondary outcomes included fluid overload events, development of shock requiring intravenous boluses, development of neurological complications; severe electrolyte abnormalities at 24 hours and day 28 mortality.

**Findings:** We identified 3 RCTs, comprising 484 participants with severe malnutrition including 72 children with the kwashiorkor phenotype: (2 some risk of bias; 1 low risk of bias). The risk ratio (RR) for in-hospital mortality with IVR versus OR was 0.71 (95% confidence interval [CI], 0.46-1.10; I^2^=0.0%) with moderate certainty of evidence. No fluid overload events were reported, pooled RR 0.99 (95% CI 0.10-9.35). Pooled RR of severe hyponatremia at 24 hours (grouped by threshold (sodium < 125 or <130mmol/L)) was 0.66 (95% CI 0.44-0.99). Only one trial reported RR for shock development; hypernatremia (sodium >145mmol/l) and 28-day mortality with IVR versus OR RRs of 0.56 (95% CI 0.21-1.48); 2.05 (95% CI 0.50-8.58) and 0.85 (95% CI 0.44-1.65) respectively. Subgroup analyses for in-hospital mortality were carried out for region and risk of bias rating giving p=0.85 and p=0.54 for heterogeneity respectively.

**Conclusion:** The estimated effect of using IVR versus OR in children with SAM with severe dehydration ranges from a 54% relative reduction to a 10% relative increase in the risk of death with IVR resulting in fewer adverse events.

(PROSPERO number, CRD42025637956.)

**Key Questions:** *What is already known on this topic:* International guidelines advise against giving intravenous rehydration to children with severe acute malnutrition due to concerns about fluid overload. Evidence to support this recommendation is weak, specifically in children in African with severe dehydration due to diarrhoea

*What this study adds:* We systematically reviewed evidence comprising of 484 children in 3 randomised trials, and estimated that intravenous rehydration resulted in a 54% relative reduction to a 10% relative increase in the risk of death No evidence of fluid overload or cardiac failure was reported in any trial

*How might this study affect research, practice or policy:* The guidance on intravenous rehydration should be reconsidered in light of the findings of this review and considering the safety of intravenous rehydration Simplification of the rehydration guidelines for severe dehydration to remove the distinction between malnourished and non-malnourished children would facilitate ease of implementation.

## Introduction

International recommendations for inpatient management of children with severe acute malnutrition (SAM) advise against the use of intravenous rehydration (IVR) for the treatment of severe dehydration. This recommendation is based on concerns about children with SAM being at high risk of cardiac compromise and sodium overload^1,2^ albeit based on limited evidence. Instead, oral rehydration (OR) is the recommended option, with intravenous (IV) boluses only to be provided in the event of shock. Whilst physiological studies have supported the safety of intravenous fluids in SAM and found no evidence of cardiac compromise^3,4^, including appropriate adaptative responses^3,5^, controlled clinical trials assessing the safety and efficacy of liberal intravenous rehydration are lacking^6^.

Dehydration due to gastroenteritis is a common complication in children with SAM and remains an important cause of morbidity and hospitalization associated with poor outcomes reported^7–9^-suggesting that the current standard management might be suboptimal. The recent results from GASTROSAM (<u>G</u>astroenteritis: <u>R</u>ehydration <u>o</u>f children with <u>S</u>evere <u>A</u>cute Malnutrition)^10^, the largest trial to date conducted in Africa addressing this question, justifies a comprehensive re-examination of the evidence.

Our objective was to systematically review the literature and assess the efficacy and safety of intravenous rehydration (IVR) compared to standard care (oral rehydration; OR) in children hospitalized with SAM complicated by severe dehydration due to gastroenteritis.

## Methods

This systematic review and meta-analysis were conducted following the PRISMA (Preferred Reporting Items for Systematic Reviews and Meta-Analyses) recommendations (**Supplementary Table S1**). The review protocol was submitted to the PROSPERO repository (CRD42025637956) before the review was conducted.

### Eligibility Criteria

We included randomized controlled trials (RCT) and quasi-randomized controlled trials evaluating children aged 5 months to 12 years hospitalized with severe malnutrition (defined as any one of: weight-for-height Z-score less than −3 or mid-upper arm circumference less than 11.5cm or oedematous malnutrition (kwashiorkor: bilateral pedal oedema or more generalized oedema) with gastroenteritis/diarrhea (> 3 loose stools/day) and severe dehydration (following WHO criteria, include two or more of the following: altered consciousness (less than Alert on AVPU score), sunken eyes, reduced skin turgor (slow abdominal skin pinch return>2 seconds) or unable to take or retain oral fluids). The trials compared, by intention, one group rehydrated intravenously (IVR) with fluids of any type and volume (intervention group) to an oral rehydration (OR) strategy (defined as the control). As per WHO guidelines the control group were permitted to receive IV fluids to treat hypovolemic shock (defined as any of capillary refill> 3 seconds, weak pulse, cool peripheries); oral rehydration was permitted in all children (as a follow on from IVR for children with ongoing losses). We planned to exclude studies which enrolled children with known uncorrected congenital heart abnormality or pre-existing renal conditions.

### Information Sources

MEDLINE, Embase, CINAHL, and Cochrane Central Register of Controlled Trials (CENTRAL, The Cochrane Library) were searched. We searched for relevant references cited in the papers identified through the above databases. We also searched for ongoing clinical trials though ClinicalTrials.gov, ISRCTN registry, and the World Health Organization International Clinical Trials Registry Platform to include potentially validated data in the process of being published but not yet accessible. The last searches were conducted on 15 March 2025.

### Search Strategy and Selection Criteria

The search strategy was based on: IV fluid management, children, severe acute malnutrition, dehydration, and diarrhea (or gastroenteritis). We listed key search terms, their synonyms, and Medical Subject Headings (MeSH) for each concept. We then conducted the search combining the key search terms, their synonyms, and MeSH with appropriate Boolean operators (Supplementary: **Search Strategies Table**). No time or language restrictions were applied.

Search results were imported into a references management software (Zotero®). Duplicates were removed through a specific feature of the software. Paired reviewers (MC, SO, HAS, TS) independently conducted title and abstract screening (including full-text screening when titles and abstracts provided insufficient information). MC and SO screened half of the articles, HAS and TS screened the other half. Discrepancies were resolved through consensus by discussion with an additional reviewer (JED). Screening was based on the eligibility criteria described above.

### Data Extraction

Data were extracted on publication characteristics, country, baseline demographics including SAM definition, dehydration definition, study design, inclusion criteria, exclusion criteria, description of intervention, description of control care, number of participants assigned to each group, and the predefined outcomes of each study and reported subgroup analyses. Articles not found to meet the study inclusion criteria were excluded, with the reasons recorded. A standardized data extraction form was used to extract data needed to describe the included articles, assess their quality, and perform the metanalysis.

Data was extracted by the paired reviewers. Discrepancies were resolved through consensus by discussion with an additional reviewer (JED).

### Outcomes and subgroup analysis

The prespecified primary outcome was in-hospital mortality. Secondary outcomes were fluid overload events (pulmonary oedema or heart failure), development of shock requiring intravenous boluses, development of neurological complications (convulsions or decrease in conscious level); presence of severe electrolyte abnormalities (hyponatremia, hypernatremia or hypokalemia as defined by the study), day-28 mortality, and hospital readmission. Prespecified subgroup analyses included for the primary outcome (in-hospital mortality) compared trials in Africa vs Asia; children under 1 year and presence of kwashiorkor.

### Risk-of-Bias Assessment

Paired reviewers (JED, CM, TS) conducted risk-of bias assessments independently (JED assessed all papers, and CM, TS each assesses one half of the papers independently) using the Risk of Bias 2.0 tool^11^. Discrepancies were solved through consensus discussions between JED, CM, and TS. Risk-of-bias was classified as low, high, or some concerns for the primary outcome (mortality before discharge). The assessment was based on randomization process, deviations from the intended intervention, missing outcome data, measurement of the outcome, and selection of the reported result, as per the Risk of Bias 2.0 tool instructions.

### Statistical Analyses

Meta-analyses were performed in Stata v18.0 with random-effects and using restricted maximum likelihood. All outcomes are presented as risk ratios; only outcomes reported in two or more studies were analyzed. Heterogeneity between studies was assessed using I^2^ and visual inspection of forest plots. One planned subgroup analysis was conducted by region and a post-hoc sub-group analysis by risk of bias was performed as part of the GRADE assessment. Other subgroups based on age and presence of kwashiorkor (oedematous malnutrition) could not be analyzed as the specific data were not disaggregated in the published reports.

### Certainty of Evidence

The overall certainty in evidence for each outcome of interest summarized with meta-analysis was rated using the GRADE (Grading of Recommendations Assessment, Development, and Evaluation) framework, based on risk-of-bias, imprecision, inconsistency, indirectness, and publication bias. The certainty of evidence was rated as very low, low, moderate or high^12^.

## Results

### Study Selection

Of 393 unique citations identified by the search six studies^13–18^ were assessed for potential eligibility after the initial screening. One study was excluded because it was a non-randomized observational cohort study reporting before-and-after implementation of a therapeutic bundle^13^. A second study did not include a control group and was therefore excluded^14^. A third study was excluded as it was a sequential intervention study design^15^ (see Supplemental Tables S1) Of the three randomized trials only one trial^16^ fulfilled the strict criteria outlined in the search strategy (PROSPERO CRD42025637956). Two other trials^17,18^ examined a more liberal intravenous rehydration strategy to a more conservative strategy so were included in the meta-analysis for completeness. One trial study recruited SAM children with severe dehydration or septic shock^17^. Only data for children with severe dehydration were used in analysis. Another trial included children a subset of SAM with within the studypopulation^18^. Outcomes for the SAM subset only are included in the analysis. These three studies involved 484 children in total were included in the analysis (**Figure 1 and Table 1**).

**Table 1.** Characteristics of studies included.

| Author, year, country | Study design | Participants characteristics | Description of intervention | Description of control care | Participants in intervention group | Participants in control group | Mortality before hospital discharge |
| --- | --- | --- | --- | --- | --- | --- | --- |
| Akech, 2010, Kenya | Phase II randomized controlled trial | 61 children over 6 months of age were enrolled. Forty children had hypovolaemic shock secondary to dehydrating diarrhoea (an additional 21 had hypovolaemic shock secondary to sepsis and are not included in this analysis) | Ringers Lactate (RL) resuscitation: initial bolus of 10 ml/kg over 30 minutes, repeated only twice over one hour (i.e. up to 30 ml/kg in total). Additional boluses (10 ml/kg over one hour) were only permitted if oliguria or hypotension. Maximum bolus volumes given were 40 ml/kg.<br>ReSoMal (oral Rehydration Solution for <u>Ma</u>lnutrition) was given to children with significant diarrhoea | Initial bolus of 15 ml/kg of half-strength Darrow's in 5% dextrose (HSD/5D) over one hour. Repeat bolus was given once (15 ml/kg of HSD/5D over 1 hour) if some improvement in features of shock noted. If no improvement was seen, they received 10 ml/kg whole blood transfusion over 3 hours.<br>ReSoMal was given to children with significant diarrhoea | 21 | 19 | Overall, 22/40 children died.<br>Intervention: 9/21<br>Control: 13/19 |
| Alam, 2020, Bangladesh | Open randomized controlled clinical trial | Malnourished children aged 6-60 months, with a mixed population of severe acute malnutrition (172) and chronic malnutrition (28) | Isotonic IV solution at a rate of 100 ml/kg over six hours. Intravenous fluid infusion was stopped after six hours, and the ongoing stool loss was replaced with a rice-based ORS and continued until the diarrhoea resolved. | Initially, children received 15 mL/kg of an isotonic IV solution over one hour. If their respiratory and pulse rate slowed down after one hour, the intravenous fluid was continued at 15 mL/kg for another hour. After two hours, intravenous fluids were stopped, and a rice-based ORS was provided at 5-10 ml/kg to provide a total of 70 ml/kg of ORS. | 105 (85 SAM) | 103 (87 SAM) | 0/85 in intervention arm<br>0/87 in control arm |
| Maitland, 2025, Kenya, Uganda, Niger, Nigeria | Open label randomized controlled clinical trial | Severely malnourished children aged 6 months to 12 years with gastroenteritis and severe dehydration with or without hypovolaemic shock | Children were randomised between two intervention strategies (then combined for analysis as a liberal strategy). 1) rapid IV rehydration: 100 ml/kg RL over 3-6 hours according to age including boluses (20 ml/kg) for those with shock) 2) A slower IV rehydration regimen (100 ml/kg RL given over 8 hours and no boluses) | ORS (second randomisation between standard WHO ORS (usually given for non-SAM children) or ReSoMal at a rate of 5 ml/kg every 30 min for the first 2 h followed by 5–10 ml/kg per h for the next 4–10 h on alternate hours, with F-75 milk nutrition formula plus boluses (15ml/kg) given for shock. Bolus of Ringer's lactate given over 1 hour and repeated if necessary. | 134 | 138 | 13/134 in intervention; 16/138 in control |

**Figure 1.**
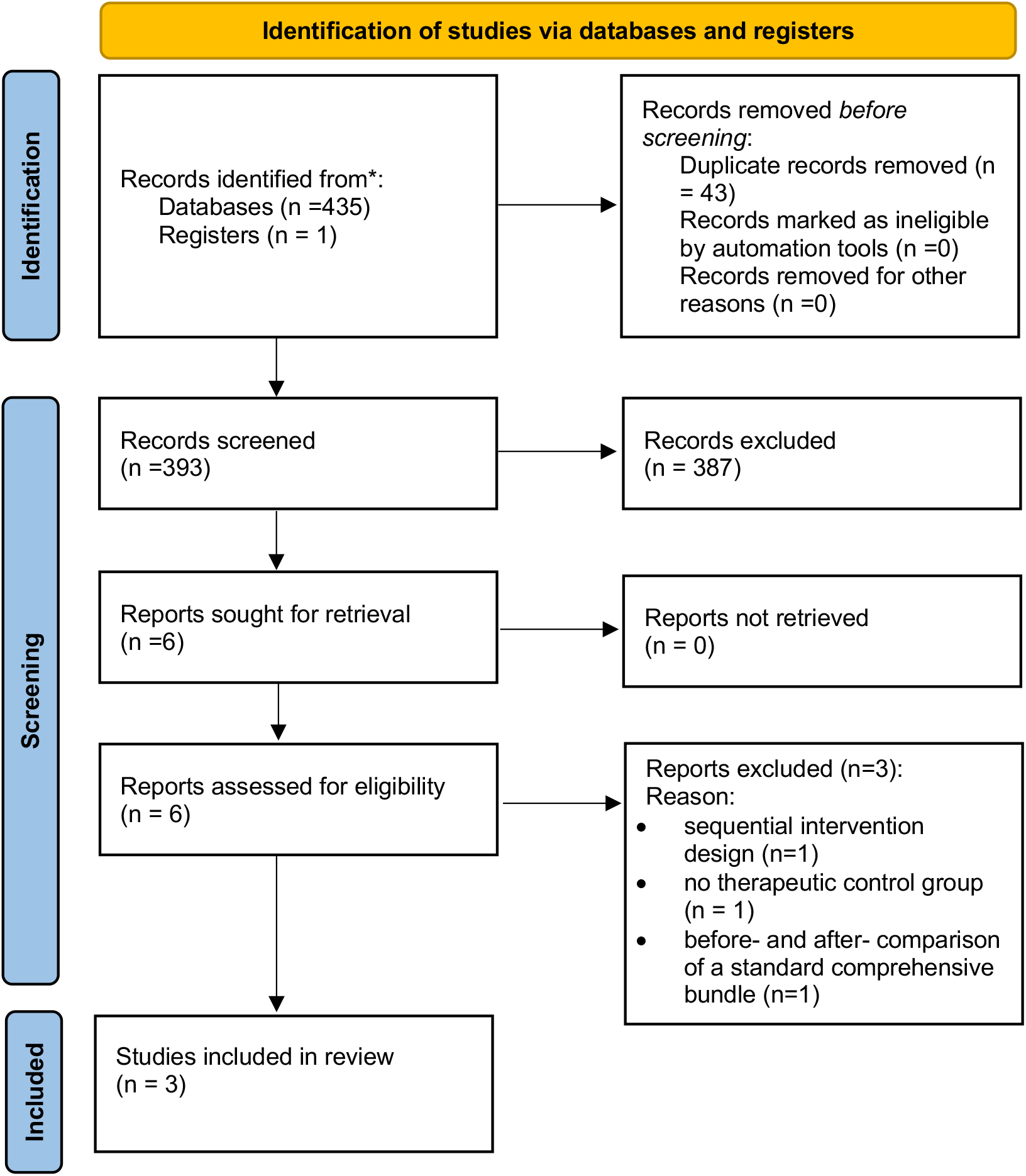
PRISMA Flow diagram.

### Study Characteristics

The three included studies were a multinational RCT, conducted in Kenya, Uganda, Niger, and Nigeria^16^; a RCT conducted in Kenya^17^ and one RCT conducted in Bangladesh^18^ (**Table 1 and Tables S2)**. The first RCT included children between 6 months and 12 years (median age 13 months)^16^. In one study children >6 months (median ages of 15 and 16 months) in the intervention and control arms respectively were included^17^ and one study included children aged 6 to 60 months^18^. This RCT included children with SAM and chronic malnutrition^18^, however the other two studies included only children with SAM^16,17^. The kwashiorkor phenotype of severe malnutrition was present in 11 (4%)^16^, 12 (22%)^17^ and 49 (24%)^18^ of study participants. All three trials included children with severe dehydration. In two studies children presented with diarrhea and reported the numbers with invasive bacterial infection(bacteraemia)^16,17^ were reported, while one study included only children with diarrhea^18^.One RCT enrolled children with severe dehydration and hypovolemia shock (40/61 participants) and septic shock without dehydration (21/61 participants)^17^. The data from the latter group was excluded in this systematic review. The RCT from Bangladesh compared the provision of 100ml/kg of an isotonic IV solution over 6 hours (intervention) versus up to 30 ml/kg given over 2 hours (same solution) followed by an OR (control)^18^.. The multicentre RCT evaluated three interventions: immediate rapid rehydration (100 ml/kg RL over 3-6 hours according to age), immediate slow rehydration (100 ml/kg RL given over 8 hours) (both considered as intervention), versus control (oral rehydration solution at a rate of 5 ml/kg every 30 min for the first 2 h followed by 5–10 ml/kg per h for 4–10 h). No intravenous rehydration IVR was permitted in the control arm, per protocol fluid boluses were reserved for with or developing shock^16^. The RCT in Kenya compared providing IV boluses of Ringer’s Lactate (RL) (up to 40 ml/kg) (intervention) versus 1-2 IV boluses of 15 ml/kg intravenous half-strength Darrow’s/5% dextrose (HSD/5D) (control). Oral Rehydration <u>So</u>lution for <u>Mal</u>nutrition (ReSoMal) was given to children with significant diarrhea in both arms^17^.(see **Table S2b**)

### Risk of Bias in Studies

Overall, two studies^17,18^ were assessed as presenting some concerns on risk of bias domains, and one as having a low risk of bias^16^. In the two studies with some concern, we estimated that there was a risk in the selection of the reported results which may have resulted in bias (**Figure 2a**).

**Figure 2a.**
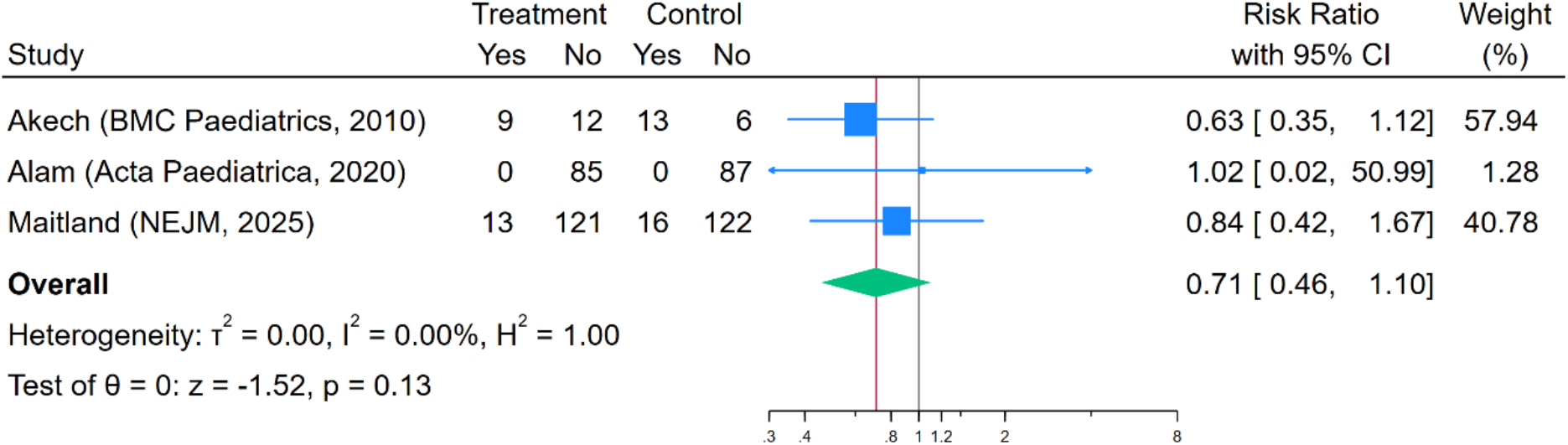
Forest plot of in-hospital mortality.

### Primary Outcome

The pooled estimated risk ratio (RR) for in hospital mortality was 0.71 (95% confidence interval (CI) 0.46, 1.1) I^2^ = 0.0% (**Figure 2a; Table 2 and Table S3**). This translates to an estimated effect of using IVR versus OR in children with SAM with severe dehydration ranges from a 54% relative reduction to a 10% relative increase in the risk of in-hospital mortality. The GRADE summary indicates moderate certainty of evidence for the primary outcome, with no evidence of a difference between intervention and control strategies for rehydration of children with severe acute malnutrition and severe dehydration.

**Table 2.** Evidence profile for outcomes from meta-analysis.

| Outcome | Meta-analysis results, Risk Ratio (95%CI) | Certainty of evidence | Risk of bias | Inconsistency | Indirectness | Imprecision | Publication bias |
| --- | --- | --- | --- | --- | --- | --- | --- |
| In-hospital mortality | 0.71 (0.46, 1.10) | Moderate due to serious imprecision. | Results from trials with some concern and low risk are concordant so no rating down | Visual exploration of forest plot and heterogeneity statistic $I^2=0\%$ indicated no inconsistency. | The populations and interventions were deemed similar enough that no rating down was needed | 95%CI of overall effect estimate crosses null so rating down one level for imprecision | Visual examination of the funnel plot does not suggest publication bias so no rating down was needed |
| Heart failure or fluid overload events | 0.99 (0.1, 9.35) | Moderate, due to serious imprecision. | Results from trials with some concern and low risk are concordant so no rating down | Visual exploration of forest plot and heterogeneity statistic $I^2=0\%$ indicated no inconsistency. | The populations and interventions were deemed similar enough that no rating down was needed | 95%CI of overall effect estimate crosses null so rating down one level for imprecision | Visual examination of the funnel plot does not suggest publication bias so no rating down was needed |
| Hyponatraemia at 24 hours | 0.66 (0.44,0.99) | Moderate, due to serious inconsistency. | Results from trials with some concern and low risk are concordant so no rating down | Visual exploration of forest plot and heterogeneity statistic $I^2=0\%$ indicated no inconsistency. However, definitions were not consistent between studies so rating down one. | The populations and interventions were deemed similar enough that no rating down was needed | 95%CI of overall effect estimate does not cross null so no rating down was needed | Visual examination of the funnel plot does not suggest publication bias so no rating down was needed |
| Hypokalaemia at 24 hours | 1.16 (0.93, 1.46) | Low, due to serious imprecision and inconsistency. | Results from trials with some concern and low risk are concordant so no rating down | Visual exploration of forest plot and heterogeneity statistic $I^2=0\%$ indicated no inconsistency. However, definitions were not consistent between studies so rating down one. | The populations and interventions were deemed similar enough that no rating down was needed | 95%CI of overall effect estimate crosses null so rating down one level for imprecision | Visual examination of the funnel plot does not suggest publication bias so no rating down was needed |

### Secondary outcomes

No fluid overload events were reported in any RCT, the pooled RR for fluid overload in IVR versus OR was 0.99 (95% CI 0.10, 9.35) (**Figure 2b; Table 2**) with moderate certainty of evidence. Severe hyponatremia and hypokalemia at 24 hours, was reported in two studies, with different thresholds (as <125mmol/L and <2.5mmol/L respectively^16^ and <130 and <3.5 respectively^18^). The different thresholds were defined as subgroups, and the pooled estimated RR was 0.66 (95% CI 0.44, 0.99) for hyponatremia (**Supplemental Figure S1a**) with moderate certainty of evidence and 1.16 (95% CI 0.93, 1.46) with low certainty of evidence for hypokalemia (**Supplemental Figure S1b)**.

**Figure 2b.**
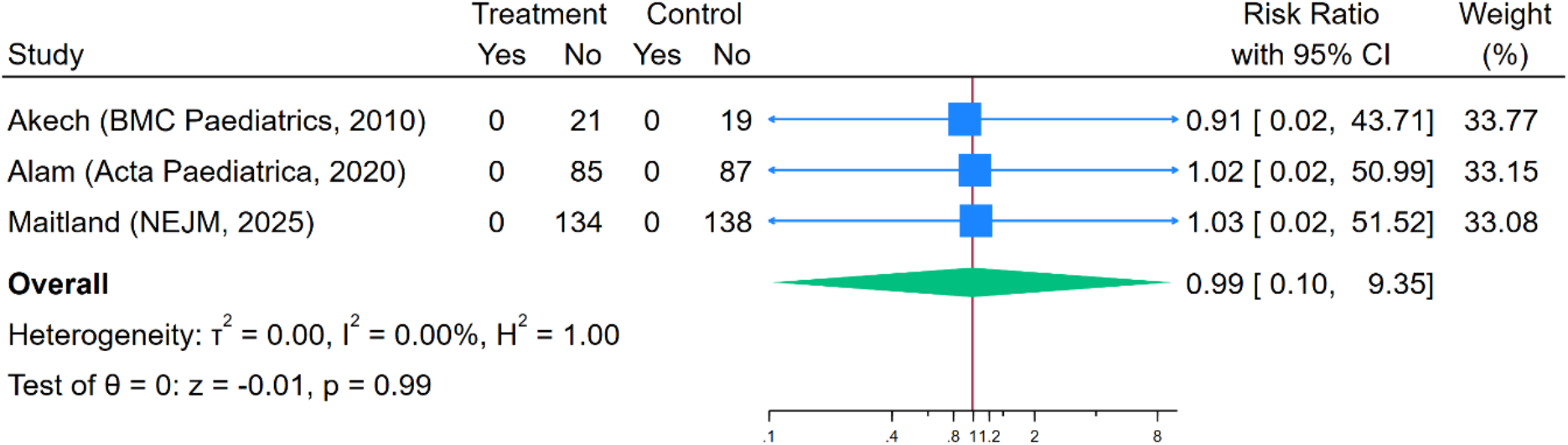
Forest plot of heart failure of fluid overload events.

**Figure 3.**
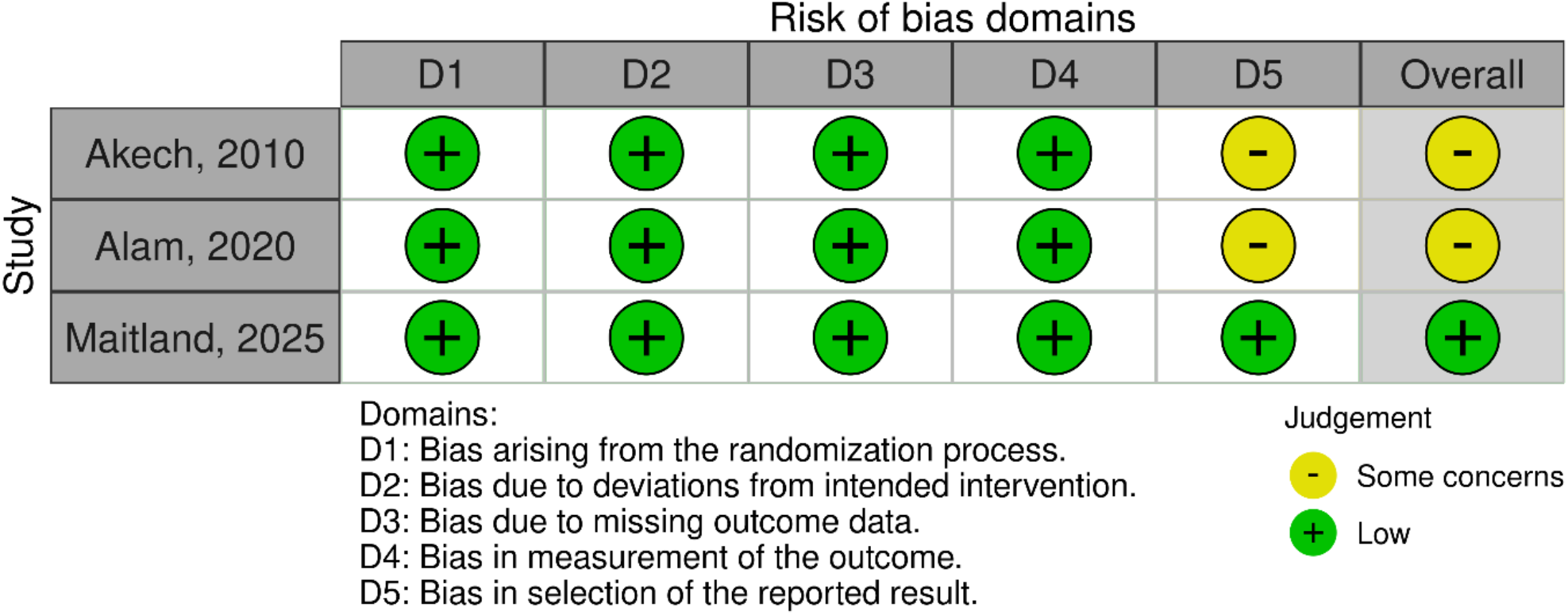
Risk of Bias plot.

Other pre-specified outcomes were only reported in one RCT^16^, so were not pooled for analysis, including: development of shock, development of neurological complications (convulsions or decrease in conscious level) during the primary admission; day 28 mortality and severe hypernatremia (> 145 mmol/L). In that trial for the IVR versus OR comparison for shock development, occurring in 6/134 vs 11/136 respectively the RR was 0.56 (95% CI 0.21-1.48); hypernatremia (sodium >145mmol/l) occurred in 6/127 vs 3/128 respectively with RR 2.05 (95% CI 0.50, 8.58). Day 28 mortality was reported in 14/134 vs 17/138 respectively with RR 0.85 (95% CI 0.44, 1.65). Neurological complications were reported in one participant (in the IVR arm)^16^. Readmissions were not reported in any study. **(Table S2a and S2b)**

### Sub-group Analyses

Subgroup analysis for Africa and Asia for the primary outcome of mortality demonstrated a pooled estimated risk ratio of 0.71 (95% CI 0.46, 1.10) (**Supplemental Figure S2**) with no evidence of heterogeneity between the regions. In-hospital mortality in the subgroups of children < 12 months of age or kwashiorkor were not reported as the data were not disaggregated in more than one trial^17,18^.

### Additional Analyses

Visual examination of the funnel plot for in-hospital mortality (**Figure S2**) did not suggest publication bias. There was no evidence of heterogeneity between studies with low risk and some concern (p=0.54; **Figure S3**).

## Discussion

Our search strategy only yielded one trial which strictly fulfilled our critieria for comparing a liberal intravenous rehydration versus a control oral strategy. For completeness we included two RCT in which the rehydration strategies were either more or less conservative with regard to intravenous rehydration.

The systematic review, involving three trials, found a moderate certainty of evidence indicating the estimated effect of using IVR versus OR in children with SAM with severe dehydration ranges from a 54% relative reduction to a 10% relative increase in the risk of in-hospital death. No event of fluid overload was reported in any of the RCTs. Also of relevance, no events of fluid overload were reported in the 2 excluded studies involving 169 children receiving IVR^14,15^. The estimated pooled estimate of severe hyponatremia at 24 hours (per study definitions) in IVR versus OR strategies ranged from 66% to a 1% relative reduction.

Although formal evidence to support or refute superiority of IVR remain limited our systematic review underscores the paucity of data generated in clinical trials with only one incorporating a control strategy reflecting the current recommended rehydration guideline. Thus, reflects the limited internal validity of the results for both the primary outcome (albeit indicating some benefit associated with intravenous rehydration). This highlights the major challenges of conducting high quality research under extremely challenging conditions and the perceived ethics of conducting such trials.

A large observational cohort study was excluded (see Tables S1 for details) from this systematic review which is frequently cited in support of current rehydration guidelines for SAM^19,20^, apparently showing lower mortality with less IV rehydration use. The study reported in 1999 involved 627 children hospitalised with SAM in Bangladesh, comparing 293 admitted pre- and 334 post-implementation of a new therapeutic treatment bundle. At baseline, only 40% (138 and 117 respectively) had any signs of dehydration. The new treatment bundle included less IV rehydration in addition to several other elements of care (systematic antibiotics and micronutrients, prevention of hypoglycaemia, training of staff and availability of standardised protocols) which were not provided in the pre-implementation cohort. Moreover, the study did not reflect the current guidance for exclusive oral rehydration management despite it being a key reference source.

## Limitations

A key limitation is that from the available data only 3 trials were eligible for inclusion including 484 children in total and all of our prespecified secondary endpoints were reported in all the RCT. Overall, there was insufficient evidence to indicate whether IVR results in either superior or inferior efficacy (in-hospital mortality) than those managed on the conservative oral rehydration protocol. However, with respect to safety, the combined data from the RCTs included in the meta-analysis found no evidence of adverse events indicating fluid overload with IVR. A subgroup analysis for kwashiorkor or for children under one year were not possible since at least one trial did not report data for these groups. However, taken together, the data from the all the available studies (reporting fluid overload) who received liberal IVR (either slow or fast) indicates that the overall finding of no evidence of fluid overload (including 72 children with kwashiorkor who received IV rehydration/volume correction) that the findings of this meta-analysis are likely to extend to these sub-groups

## Implications for practice

This systematic review provides further evidence that concerns about fluid overload in SAM children receiving IVR are unfounded. When considered alongside of earlier physiological studies, showing no evidence of cardiac dysfunction, including subgroup of children with kwashiorkor ^3,4^, whose Frank-Starling curves during rehydration demonstrated preserved fluid-responsiveness^3^, these findings provide compelling evidence against the commonly held fears over use of IVR intravenous fluids for children with SAM. The GASTROSAM RCT was instigated to address these specific safety concerns and explored two IVR strategies. Indeed, for the current oral rehydration recommendations, where a child is expected to drink almost 10% of their body weight over 10 hours, in practice we found in the control OR arm that at admission 79% of children were unable to take oral rehydration, resulting in 92% requiring this via a nasogastric tube. Thus, current recommendations led to additional demands on nurses since OR could not be given by the child’s caregiver. Although overall mortality (9%) was substantially lower than the 58% predicted from similar cohorts, we suggest this was a result of the close clinical monitoring afforded by the trial for ethical reasons. Outside of a clinical trial this would not be possible in hospitals with limited clinical personell. Thus, a simplification of the rehydration protocol would potentially be easier to implement. Of note, relevant to current recommendations the product specifications for ReSoMal indicate it should not be administered unless children are ‘within an inpatient facility under strict medical supervision and under close monitoring’ for signs of fluid overload (https://supply.unicef.org/s1561125.html). Such specifications have un-intended consequences for children with SAM managed in the community with less severe degree of dehydration and may underpin the poor outcomes reported^21^. Relevant to the broader population of children hospitalised with acute diarrhoea with severe dehydration (~10% weight loss), a study showed that approximately 20% temporarily fulfilled anthropometric criteria for SAM (MUAC<11.5cm) but were ‘reclassified’ as undernourished^22^ following rehydration. Thus, through ‘slippage’ the current recommendations may have wider implications, as potentially 20% of non-SAM children could be inappropriately diagnosed as malnourished and rehydrated. This may have contributed to the poor outcomes observed in the Global Enteric Multicentre (GEMS) study^23^.

## Conclusion

A conclusion which could be drawn is that there is a need for more RCTs to establish the relevant evidence to change guidelines. However, guidelines were established, albeit on a poor evidence base, to reduce the putative risk of fluid overload from IVR. The finding of no events in the studies included in this systematic review challenges the equipoise and ethical rationale for future investigation.

## Data Availability

All data produced in the present study are available upon reasonable request to the authors

## Role of the authors

JED, TS MEC RP ECG and KM designed the study protocol; JED, MEC SO HAS TS extracted and reviewed the relevant data; CM and ECG analysed the data. KM wrote the first draft of the manuscript, which was reviewed and agreed on by all the authors.

## Availability of data and materials

The datasets analysed during the current study are available from the corresponding author on reasonable request.

## Ethics Approval Statement

Not relevant

