## Supplemental File for "Intravenous rehydration in children with severe malnutrition: a systematic review and meta-analysis"

### Supplementary material

#### Search strategy Tables

1. Medline

| # | Query |
|---|---|
| 1 | (Fluid therapy or fluid-therapy).tw. |
| 2 | ((intraven* or IV or parenteral) adj infusion).tw. |
| 3 | ((intraven* or IV or parenteral) adj administration).tw. |
| 4 | exp administration, intravenous/ or infusions, intravenous/ |
| 5 | Fluid Therapy/ |
| 6 | (infant$ or baby or babies or child$ or boy$ or girl$ or preschool$ or pre-school$).tw. |
| 7 | exp child/ or infant/ |
| 8 | (malnutrition or mal-nutrition or malnourish$ or wasted or wasting or emaciat$ or undernutrition or undernourish$ or marasmus$ or kwashiorkor).tw. |
| 9 | exp Severe Acute Malnutrition/ |
| 10 | Protein-Energy Malnutrition/ |
| 11 | (dehydration or hypovolemia).tw. |
| 12 | exp Dehydration/ |
| 13 | (diarrh$ or gastroenteritis or gastrointestinal infection$ or enteritis).tw. |
| 14 | exp Diarrhea/ |
| 15 | 1 or 2 or 3 or 4 or 5 |
| 16 | 6 or 7 |
| 17 | 8 or 9 or 10 |
| 18 | 11 or 12 |
| 19 | 13 or 14 |
| 20 | 15 and 16 and 17 and 18 and 19 |

2. Embase

| # | **Query** |
|---|---|
| 1 | (Fluid therapy or fluid-therapy).tw. |
| 2 | ((intraven* or IV or parenteral) adj infusion).tw. |
| 3 | ((intraven* or IV or parenteral) adj administration).tw. |
| 4 | exp intravenous drug administration/ |
| 5 | exp fluid therapy/ |
| 6 | (infant$ or baby or babies or child$ or boy$ or girl$ or preschool$ or pre-school$).tw. |
| 7 | exp child/ or exp infant/ |
| 8 | (malnutrition or mal-nutrition or malnourish$ or wasted or wasting or emaciat$ or undernutrition or undernourish$ or marasmus$ or kwashiorkor).tw. |
| 9 | exp malnutrition/ |
| 10 | exp protein calorie malnutrition/ |
| 11 | exp wasting syndrome/ or exp kwashiorkor/ or exp marasmus/ |
| 12 | (dehydration or hypovolemia).tw. |
| 13 | exp dehydration/ |
| 14 | (diarrh$ or gastroenteritis or gastrointestinal infection$ or enteritis).tw. |
| 15 | exp diarrhea/ |
| 16 | 1 or 2 or 3 or 4 or 5 |
| 17 | 6 or 7 |
| 18 | 8 or 9 or 10 or 11 |
| 19 | 12 or 13 |
| 20 | 14 or 15 |
| 21 | 16 and 17 and 18 and 19 and 20 |

3. CINAHL

| | | |
|---|---|---|
| S19 | S14 AND S15 AND S16 AND S17 AND S18 | Expanders - Apply equivalent subjects<br><br>Search modes - Proximity |
| S18 | S12 OR S13 | Expanders - Apply equivalent subjects<br><br>Search modes - Proximity |
| S17 | S10 OR S11 | Expanders - Apply equivalent subjects<br><br>Search modes - Proximity |
| S16 | S7 OR S8 OR S9 | Expanders - Apply equivalent subjects<br><br>Search modes - Proximity |
| S15 | S5 OR S6 | Expanders - Apply equivalent subjects<br><br>Search modes - Proximity |
| S14 | S1 OR S2 OR S3 OR S4 | Expanders - Apply equivalent subjects<br><br>Search modes - Proximity |
| S13 | (MH "Diarrhea") | Expanders - Apply equivalent subjects<br><br>Search modes - Proximity |
| S12 | diarrh* or gastroenteritis or gastrointestinal infection* or enteritis | Expanders - Apply equivalent subjects<br><br>Search modes - Proximity |
| S11 | (MH "Dehydration") | Expanders - Apply equivalent subjects<br><br>Search modes - Proximity |
| S10 | dehydration or hypovolemia | Expanders - Apply equivalent subjects<br><br>Search modes - Proximity |
| S9 | (MH "Wasting Syndrome+") | Expanders - Apply equivalent subjects<br><br>Search modes - Proximity |
| S8 | (MH "Malnutrition+") OR (MH "Protein-Energy Malnutrition+") OR (MH "Kwashiorkor") | Expanders - Apply equivalent subjects<br><br>Search modes - Proximity |

| | | |
|---|---|---|
| S7 | malnutrition or mal-nutrition or malnourish* or wasted or wasting or emaciate* or undernutrition or undernourish* or marasmus* or kwashiorkor | Expanders - Apply equivalent subjects<br><br>Search modes - Proximity |
| S6 | (MH "Infant+") OR (MH "Child+") | Expanders - Apply equivalent subjects<br><br>Search modes - Proximity |
| S5 | infant* or baby or babies or child* or boy* or girl* or preschool* or pre-school* | Expanders - Apply equivalent subjects<br><br>Search modes - Proximity |
| S4 | (MH "Administration, Intravenous+") OR (MH "Intravenous Therapy+") OR (MH "Infusions, Intravenous") | Expanders - Apply equivalent subjects<br><br>Search modes - Proximity |
| S3 | (intraven* or IV or parenteral) W1 administration | Expanders - Apply equivalent subjects<br><br>Search modes - Proximity |
| S2 | (intraven* or IV or parenteral) W1 infusion | Expanders - Apply equivalent subjects<br><br>Search modes - Proximity |
| S1 | “Fluid therapy” or fluid-therapy | Expanders - Apply equivalent subjects<br><br>Search modes - Proximity |

4. Cochrane Library

| | **Search:** |
|---|---|
| #1 | ("Fluid therapy" or "fluid-therapy" or intraven* NEAR infusion or IV NEAR infusion or parenteral NEAR infusion or intraven* NEAR administration or IV NEAR administration or parenteral NEAR administration):ti,ab,kw (Word variations have been searched) |
| #2 | MeSH descriptor: [Administration, Intravenous] this term only |
| #3 | (infant* or baby or babies or child* or boy* or girl* or preschool* or pre-school*):ti,ab,kw (Word variations have been searched) |
| #4 | MeSH descriptor: [Child] explode all trees |
| #5 | MeSH descriptor: [Infant] explode all trees |
| #6 | MeSH descriptor: [Severe Acute Malnutrition] explode all trees |
| #7 | MeSH descriptor: [Dehydration] explode all trees |
| #8 | (diarrh$ or gastroenteritis or gastrointestinal infection$ or enteritis):ti,ab,kw (Word variations have been searched) |
| #9 | (malnutrition or mal-nutrition or malnourish* or mal-nourish* or wasted or wasting or emaciat* or undernutrition or under-nutrition or undernourish* or under-nourish* or marasmus* or kwashiorkor):ti,ab,kw (Word variations have been searched) |
| #10 | (dehydration or hypovolemia):ti,ab,kw (Word variations have been searched) |
| #11 | MeSH descriptor: [Diarrhea] explode all trees |
| #12 | #1 OR #2 |
| #13 | #3 OR #4 OR #5 |
| #14 | #6 OR #9 |
| #15 | #7 OR #10 |
| #16 | #8 OR #11 |
| #17 | #12 AND #13 AND #14 AND #15 AND #16 |

#### Table S1: Details of the studies excluded from the analysis

| Author, year, country | Study design | Participants characteristics | Description of intervention | Description of control care | Participants in intervention group | Participants in control group | Mortality before hospital discharge |
|---|---|---|---|---|---|---|---|
| Ahmed, 1999 Bangladesh | Before-and-after observational study | Children aged 0–5 years severely malnourished with diarrhoea with various degrees of dehydration | Patients with some dehydration: oral rehydration according to WHO guidelines Patients with severe dehydration: patients were rehydrated with 100 mL/kg of isotonic intravenous fluid infused over 6 h in infants under 12 months and over 3h in older children | Patients with some dehydration: rice-based ORS 10 mL per kg per h for the first 2 h, then 5 mL per kg per h for the next 10 h. Patients with severe dehydration: initial rehydration by isotonic IV fluid 20 mL/kg for 1 h then 10 mL/kg for 1 h. ORS 10 mL/kg/ h was started after 1 h. | 293 | 334 | 49/293 in intervention group; 30/334 in the control group |
| Alam, 2009 Bangladesh | The study was conducted in two distinct phases: 1. Phase I: safety evaluation of rapid intravenous rehydration in severely dehydrated and malnourished children through an observational study design | Children with severe malnutrition (one third of patients had acute malnutrition), aged 6 to 60 months, with acute watery diarrhoea of less than 48 hours and stool dark-field microscopy demonstrating presence of Vibrio cholerae. 149/175 patients had severe dehydration | Children with severe dehydration (149/175) were initially rehydrated with an IV ''cholera saline'' solution containing Na+ 133, K+ 13, Cl- 98, and acetate 48 (all in mmol/L) 100 mL/kg, for 4 to 6 hours. After IV rehydration, all patients (175) were randomised to 3 formulations of ORS with the same salts composition but with different substrate composition: 1) glucose- | Not applicable | 149 severely dehydrated children underwent phase 1. 175 children were then randomised to the three ORS formulations: 1) 58 to glucose-ORS arm, 2) 59 to glucose-ORS plus | Not applicable | 0/175 |

| | | | | | | | |
|---|---|---|---|---|---|---|---|
| | without a control group to assess the safety of administering rapid intravenous rehydration 2.Phase II: comparative efficacy of three ORS formulations, following the initial safety assessment, through an RCT | | ORS, 2) glucose-ORS plus 50 g/L of amylase-resistant starch, or 3) rice-ORS | | 50 g/L of amylase-resistant starch, and 3) 58 to rice-ORS | | |
| Obonyo, 2017 Kenya and Uganda | Prospective observational study | Severely malnourished Children, aged 6-60 months, with hypovolemic shock secondary to acute gastroenteritis | 10 ml/kg/h of RL over maximum 5 h. They received rehydration volume replacement without an initial fluid bolus. Children were switched to oral rehydration once they were able to tolerate oral intake or nasogastric fluids (4/9). | Bolus of 15 ml/kg of RL over 1 h, with the option of repeating it once (15 ml/kg) if signs of shock persist, followed by HSD/5D at a rate of 4 ml/kg/h. Children were switched to oral rehydration once they were able to tolerate oral intake (1/11). | 9 | 11 | 3/9 in intervention group; 5/11 in control group Day 28 mortality 5/9 vs 9/11 |

#### Table S2a: Characteristics of included studies

| **Author (Year, journal)** | **Study design** | **Country** | **SAM definition** | **Dehydration definition** | **Inclusion criteria** | **Exclusion criteria** | **Kwashiorkor** |
|---|---|---|---|---|---|---|---|
| Akech, 2010 | Phase II randomized controlled trial | Kenya | Weight for height Z (WHZ) score <-3 or weight for height percentile 70% or Mid- Upper Arm Circumference (MUAC) <11.0cm or oedema involving at least both feet | Dehydrating diarrhea defined as: ≥6 watery stools per day<br>Shock definition: WHO shock criteria were amended to include children with one or more of the following: CRT > 2 seconds, lower limb temperature gradient, weak pulse volume, prolonged capillary refill > 2 seconds, deep 'acidotic' or 'Kussmauls' breathing, creatinine >80 µmol/L, or depressed conscious state if present after correction of hypoglycemia | Children aged over 6 months with severe malnutrition with evidence of shock | 1) Severe anemia with hemoglobin less or equals to 5g/dl<br>2) Pulmonary oedema (defined as clinical evidence of presence of fine crepitations in both lung fields plus oxygen saturation <90% in air)<br>3) Raised intra-cranial pressure or known congenital heart disease | Yes, 12 patients in total |

| | | | | | | | |
|---|---|---|---|---|---|---|---|
| Alam, 2020 | Open randomized controlled clinical trial | Bangladesh | Children with weight for age (WAZ) or weight for length/height Z (WHZ) scores of less than −3, or with bipedal oedema | Severe dehydration according to a modified version of WHO guidelines | Children aged 6-60 months, severely underweight or with severe acute malnutrition, acute diarrhea (>= 3 episodes in less than 24 hours), and severe dehydration | Dysentery, severe and very severe pneumonia, suspected severe sepsis or septic shock, suspected meningitis, antibiotic use for the current illness before admission. | Yes, 49 patients in total |
| Maitland, 2025 | Open label randomized controlled clinical trial | Kenya, Uganda, Niger, Nigeria | Children with one or more of the following: MUAC <11.5cm; WHZ <-3; Kwashiorkor | Signs of dehydration as per WHO definition: two or more of decreased conscious level (AVPU<A); sunken eyes; reduced skin turgor (abdominal skin pinch goes back slowly (>2 seconds)); unable to take or retain oral fluids | Children aged 6 months to 12 years with SAM and Gastroenteritis and severe dehydration | Diarrhea lasting more than 14 days; known congenital or rheumatic heart disease; refusal of consent | Yes, 11 patients in total |

#### Table S2b: Description of the inventions given

| Author (Year, journal) | Participants age | Participants characteristics | Description of intervention | Description of control care | Number of participants in intervention group (total number and number <12 months of age) | Number of participants in control group (total number and number <12 months of age | Outcomes |
|---|---|---|---|---|---|---|---|
| Akech, 2010 | Children over 6 months | 61 were enrolled. Forty children had hypovolemic shock secondary to dehydrating diarrhea (included in the analysis) and 21 had hypovolemic shock, presumed to have been secondary to sepsis or a sepsis-like syndrome (not included in the analysis). | Ringers Lactate (RL) or albumin (HAS) resuscitation: initial bolus of 10 ml/kg over 30 minutes, repeated only twice over one hour (i.e. up to 30 ml/kg in total) if clinical reassessment demonstrated any of the features. Additional boluses (10 ml/kg over one hour) were only permitted if oliguria or hypotension. Maximum bolus volumes given were 40 ml/kg. | WHO fluid resuscitation regime: initial bolus of 15 ml/kg of half-strength Darrow's in 5% dextrose (HSD/5D) over one hour. Repeat bolus was given once (15 ml/kg of HSD/5D over 1 hour) if some improvement in features of shock noted. If no improvement was seen, they received 10 ml/kg whole blood transfusion over 3 hours. HSD/5D was an option in case of severe dehydration & shock, and in case of presumptive septic shock with no dehydration. ReSoMal (Rehydration SOlution for Malnutrition) was given to children with significant diarrhea (>6 loose stools/day) | 21 patients received RL (no data on <12 months) | 19 (no data on <12 months) | Primary outcome measurement was the resolution of features of shock at 8 and 24 hours. Secondary outcomes included incidence of adverse events and mortality |

| | | | | | | | |
|---|---|---|---|---|---|---|---|
| | | | Rehydration SOlution for Malnutrition (ReSoMal) was given to children with significant diarrhea (>6 loose stools/day) | | | | |
| Alam, 2020 | 6-60 months | Malnourished children aged 6-60 months, with a mixed population of severe acute malnutrition (172) and chronic malnutrition (28) | IV Cholera saline (sodium 133 mmol/L, chloride 98 mmol/L, potassium 13 mmol/L and bicarbonate 48 mmol/L) at a rate of 100 mL/kg over six hours. Ongoing stool losses were replaced with a rice-based oral rehydration solution that was prepared locally (5-10 | Initially, they received 15 mL/kg of the intravenous cholera saline detailed before over one hour and monitored every 10-15 minutes. If their respiratory and pulse rate slowed down after one hour, the intravenous fluid was continued at 15 ml/kg for another hour. After two hours, the intravenous fluid was stopped, and the intravenous line was left open. The rehydration process still had an estimated deficit of 70 ml/kg at this point, and the rice-based ORS was started | 105 (85 SAM) (no data on <12 months) | 103 (87 SAM) (no data on <12 months) | Administration of unscheduled IV rehydration, Treatment failure defined as not achieving rehydration after 6 hours in rapid group, after 12 hours in slow group, fluid overload, heart failure |

| | | | | | | | |
|---|---|---|---|---|---|---|---|
| | | | mL/kg was fed orally or through a nasogastric tube after each watery stool). Intravenous fluid infusion was stopped after six hours, and the ongoing stool loss was replaced with the rice-based ORS and continued until the diarrhea resolved. | through a nasogastric tube at 5-10 mL/kg. | | | |
| Maitland, 2025 | 6 months to 12 years | Severely malnourished children aged 6 months to 12 years with gastroenteritis and severe dehydration with or without hypovolemic shock | Children were randomized between two intervention strategies (then combined for analysis as a liberal strategy). 1) rapid IV rehydration as per WHO Plan C (usually for non-SAM children) (100 ml/kg Ringer's Lactate (RL) over 3-6 hours according | WHO standard of care for children with SAM and severe dehydration: ORS (second randomization between standard WHO ORS (usually given for non-SAM children) or WHO SAM-recommended low-sodium ReSoMal, at a rate of 5 ml/kg every 30 min for the first 2 h followed by 5–10 ml/kg per h for the next 4–10 h on alternate hours, with F-75 milk nutrition formula plus boluses (15ml/kg) given for shock. Bolus of Ringer's lactate | 134 | 138 | Mortality at 96hr; mortality at 28 days; fluid overload events (pulmonary edema or heart failure) during admission; change in plasma sodium levels; electrolyte abnormalities at 8 hours; weight and MUAC change to day 3 and day 7. |

| | | | | |
|---|---|---|---|---|
| | | | to age including boluses (20 ml/kg) for those with shock) 2) A slower IV rehydration regimen (100 ml/kg RL given over 8 hours and no boluses) | given over 1 hour and repeated if necessary. |

#### Table S3: Availability of the Primary and Secondary outcomes of included studies

| Author (Year, journal) | Mortality before hospital discharge | Development of pulmonary oedema or heart failure ( fluid overload) | Development of shock requiring intravenous fluids | Development of neurological complications (convulsions or decrease in conscious level) | Perturbations of electrolyte abnormalities (severe hyponatremia | Severe hypernatremia > 140 mmol/L or hypokalemia | Day-28 survival | Readmission to hospital | Were there any subgroup analyses performed |
|---|---|---|---|---|---|---|---|---|---|
| Akech, 2010 | Intervention: 9/21 Control:13/19 | Intervention group: RL: 0/21 Control group: HSD/5D: 0/19 | Not available (NA) for the subset of children with diarrhea | NA | Only baseline data was provided and then mean sodium concentrations. There were no differences in the mean sodium concentration at admission versus 8 hours, and 24 hours between intervention and control groups | Only baseline data was provided, ie electrolyte abnormalities at recruitment but no follow up data after interventions were administered | NA | NA | No |
| Alam, 2020 | 0/105 in intervention arm<br><br>0/103 in control arm | 0/105 in intervention arm<br><br>0/103 in control arm | NA | NA | 24 hours: 10/80 (12.5%) in intervention; 12/82(14.6%) in control. | Hypokalemia at 24 hours: 48/80(60%) intervention; 45/82 (54.9%) control. | NA | NA | Yes (for SAM and chronic malnutrition subgroups) |

| | | | | | | | | | |
|---|---|---|---|---|---|---|---|---|---|
| Maitland, 2025 | 13/134 in intervention; 16/138 in control | 0/134 in intervention arm; 0/138 in control arm | 6/134 in the intervention arm; 11/138 in the control arm | 10/134 in intervention arm; 18/138 in the control arm | 8hours: 20/128 (16%) in intervention; 58/126 (45%) in control.<br>24 hours: 21/127 (17%) in intervention; 35/126 (27%) in control. | Hypernatremia (only reported >145mmols/L). 8 hours: 2/128 (2%) intervention; 2/126 (2%) control. 24 hours: 6/127 (5%) intervention; 3/126 (2%) control. Hypokalemia. 8 hours 57/128 (45%) intervention; 40/126 (31%) control. 24 hours 36/127 (29%) intervention; 26/126 (20%) control. | 14/134 (10%) in intervention group; 17/138 (12%) in control. | NA | Yes (for age<1 years; conscious level (AVPU); ORS randomization; respiratory distress). |

#### Table S4: Summary of Findings table

| | Estimated risks | | | | |
|---|---|---|---|---|---|
| Outcomes | Control risks | Intervention risks | Relative effect (95% CI) | Number of participants (studies) | Quality of the evidence |
| In hospital mortality | 29/244 (11.9%) | 22/240 (9.2%) | 0.71 (0.46, 1.10) | 488 (3) | Moderate |
| Heart failure or fluid overload events | 0/244 (0%) | 0/240 (0%) | 0.99 (0.10, 9.35) | 488 (3) | Moderate |
| Hyponatraemia at 24 hours | 27/208 | 31/207 | 0.66 (0.44, 0.99) | 415 (2) | Moderate |
| Hypokalemia at 24 hours | 71/207 | 84/207 | 1.16 (0.93, 1.46) | 415 (2) | Low |

#### Figure S1a): Subgroup analysis of hyponatremia at 24 hours by definition of hyponatremia

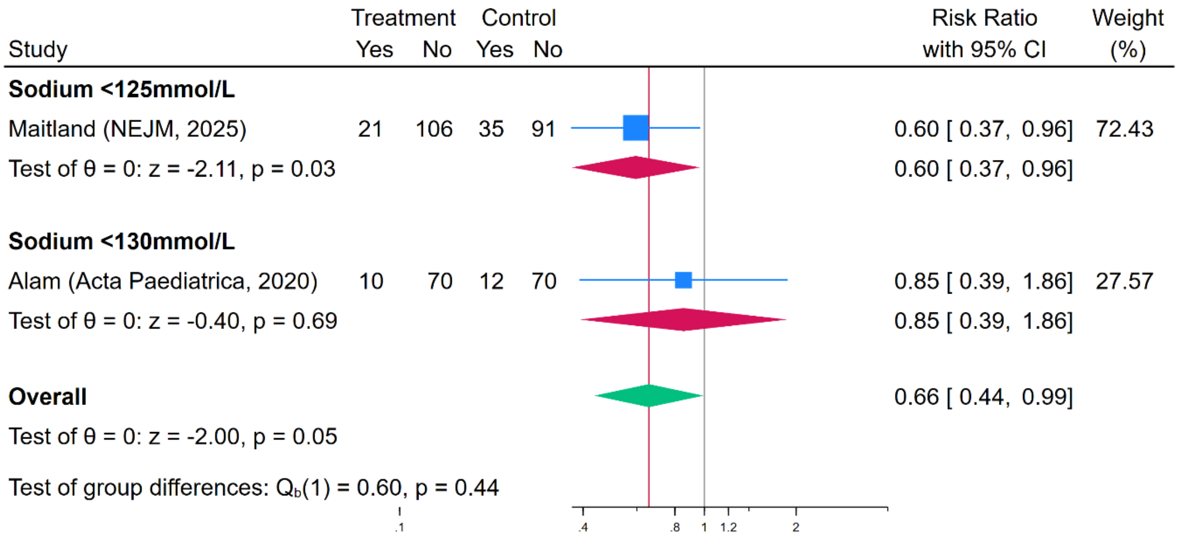

#### Figure S1b): Subgroup analysis of hypokalemia at 24 hours by definition of hypokalemia

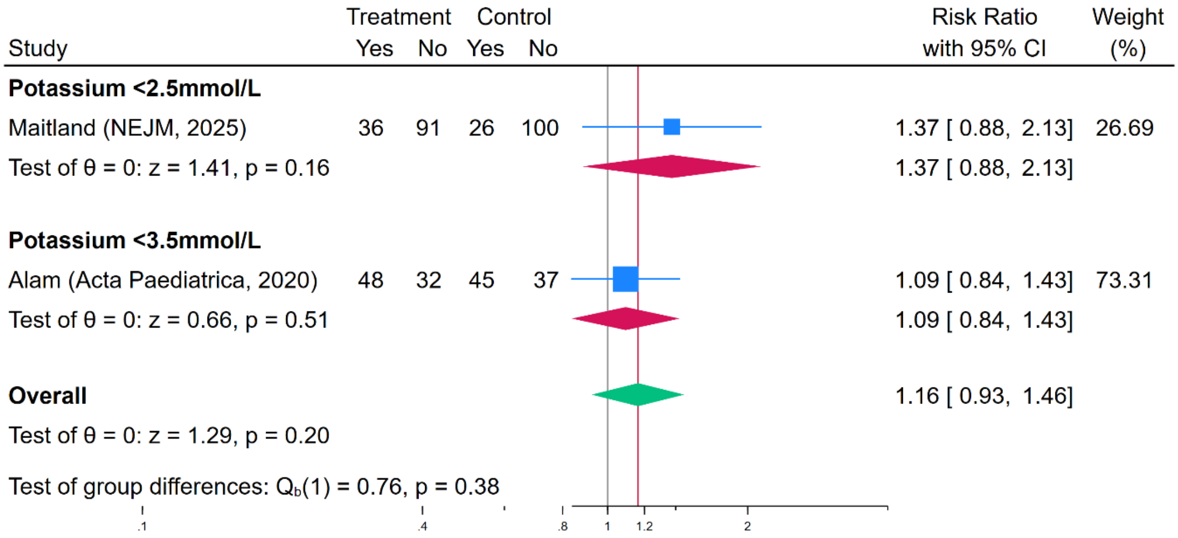

#### Figure S2: Subgroup analysis forest plot of in-hospital mortality by geographic region of study

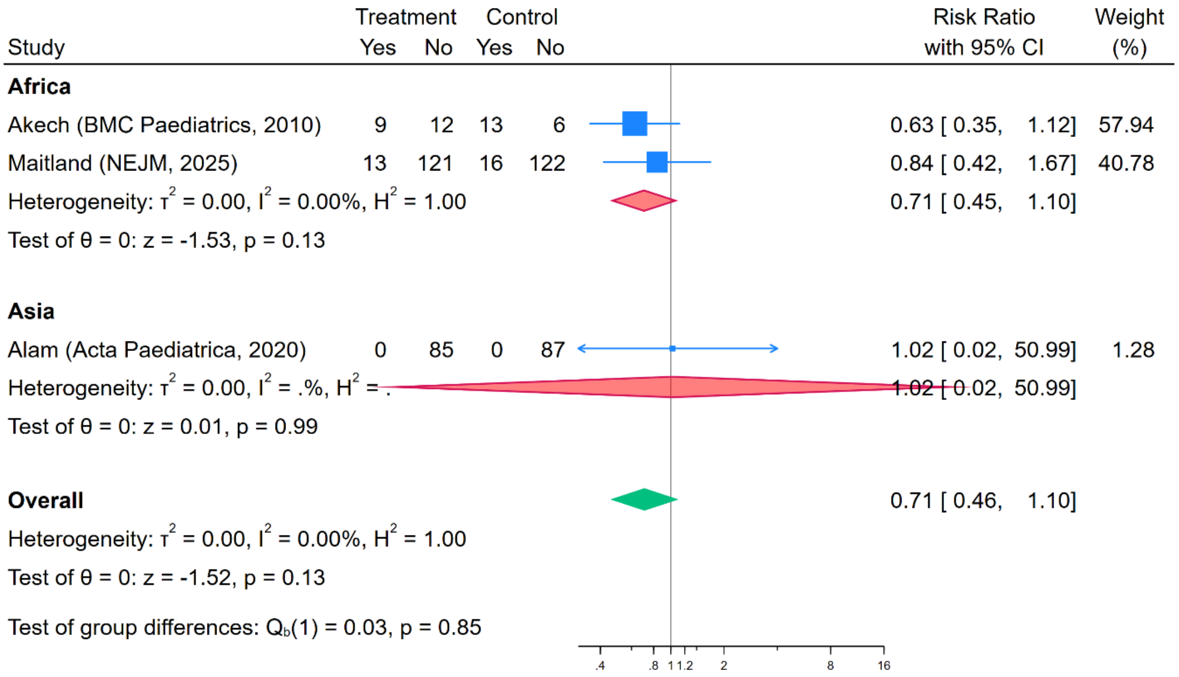

#### Figure S3: Sensitivity analysis of in-hospital mortality based on Risk of Bias assessment of study

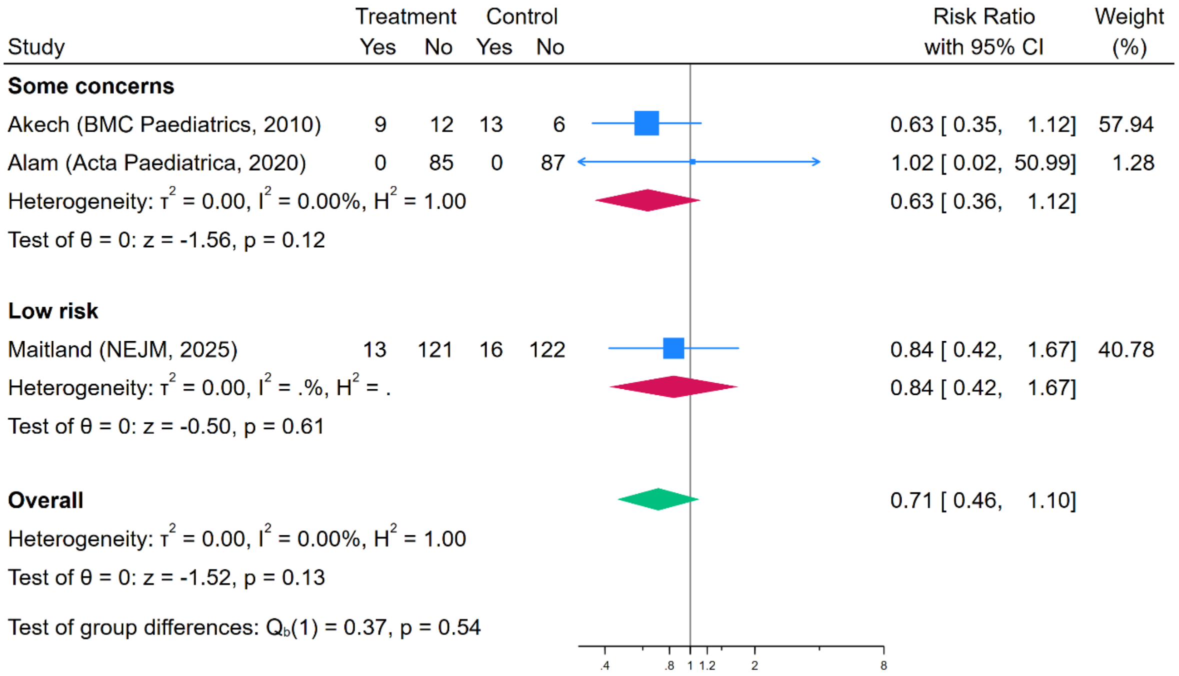

#### Figure S4: Funnel plot for mortality before hospital discharge

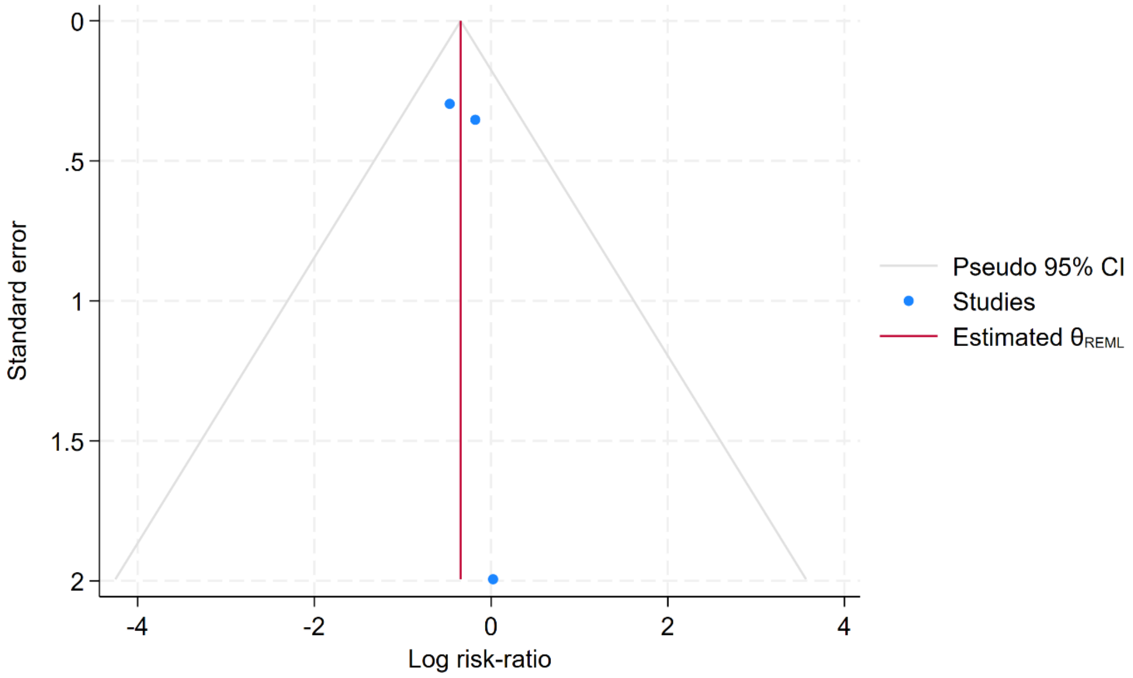
